# Joint associations of pregnancy complications and postpartum maternal renal biomarkers with severe cardiovascular morbidities: A US racially diverse prospective birth cohort study

**DOI:** 10.1101/2023.03.14.23287276

**Authors:** Xiumei Hong, Avi Z. Rosenberg, Jurgen Heymann, Teruhiko Yoshida, Sushrut S. Waikar, Titilayo O. Ilori, Guoying Wang, Heather Rebuck, Colleen Pearson, Mei-Cheng Wang, Cheryl A. Winkler, Jeffrey B. Kopp, Xiaobin Wang

## Abstract

**Rationale & Objective:** Pregnancy complications are risk factors for cardiovascular diseases (CVD). Little is known about the role of renal biomarkers measured shortly after delivery, individually or in combination with pregnancy complications, in predicting subsequent severe maternal CVD.

**Methods:** This study included 576 mothers of diverse ethnicities from the Boston Birth cohort, enrolled at delivery and followed prospectively. Plasma creatinine and cystatin C were measured 1-3 days after delivery. CVD during follow-up was defined by physician diagnoses in electronic medical records. Associations of renal biomarkers and pregnancy complications with time-to-CVD events were assessed using Cox proportional hazards models.

**Results:** During an average of 10.3±3.2 years of follow-up, 34 mothers developed one or more CVD events. Although no significant associations were found between creatinine and risk of CVD, per unit increase of cystatin C (CysC) was associated with a hazard ratio (HR) of 5.21 (95%CI = 1.49-18.2) for CVD. A borderline significant interactive effect was observed between elevated CysC (≥75th percentile) and preeclampsia. Compared to those without preeclampsia and with normal CysC level (<75^th^ percentile), mothers with preeclampsia and elevated CysC had the highest risk of CVD (HR=3.8, 95%CI = 1.4-10.2), while mothers with preeclampsia only or with elevated CysC only did not have significantly increased CVD risk. Similar synergistic effects for CVD were observed between CysC and preterm delivery.

**Conclusions:** In this sample of US, traditionally under-represented multi-ethnic high-risk mothers, elevated maternal plasma cystatin C and pregnancy complications synergistically increased risk of CVD later in life. These findings warrant further investigation.

**Clinical Perspectives:** What is new?

- Maternal postpartum elevated levels of cystatin C are independently associated with higher risk of cardiovascular diseases (CVD) later in life.
- Maternal pregnancy complications coupled with postpartum elevated levels of cystatin C synergistically increased future risk of CVD.

What are the clinical implications?

- These findings, if further confirmed, suggest that women with pregnancy complications and elevated postpartum cystatin C may be at particular high risk for CVD later in life compared to women without these risk factors.

## Introduction

Cardiovascular disease (CVD) is the leading cause of death in women worldwide. Despite declines in all other population groups, CVD mortality rates are increasing in women aged 35 to 54 years.^1^ Pregnancy is a vulnerable state. Even in a normal pregnancy, a pregnant woman experiences marked metabolic, hormonal, and physical changes to support the developing fetus.^2, 3^ These physiological challenges place pregnant women at higher risk of adverse health outcomes, compared to others. While fortunately few mothers die from pregnancy-related adverse complications, in survivors these complications are associated with increased CVD risk in later years.^4–6^ Accordingly, the American Heart Association has added a history of pregnancy complications to its list of risk factors for CVD^7^.

The physiologic mechanisms by which pregnancy-related complications contribute to future CVD are largely unexplored, while such knowledge is critically needed to predict future risk and to design effective interventions to reduce or reverse adverse trajectories. Glomerular hyperfiltration is the most notable renal adaptation to pregnancy, reflected by increasing glomerular filtration rate (GFR) and decreasing levels of serum creatinine with advancing gestational age up to 32 weeks’ gestation.^8, 9^ Understanding renal function during and after pregnancy has clinical importance, especially for those women with pregnancy complications, because impaired renal function has been associated with a subsequent burden of disease, including CVD.^10–12^

The best measure of renal function in pregnancy remains unclear. Serum creatinine is the most commonly-used measure of renal function, but during pregnancy, serum creatinine levels vary substantially as a function of gestational age, and thus, creatinine-based estimation of GFR (eGFR) may misclassify underlying renal dysfunction during pregnancy.^13^ Cystatin C (CysC) provides another useful index of renal function, with a demonstrated predictive value in acute and chronic kidney disease.^14, 15^ It has been reported to be a useful biomarker for the detection of preeclampsia.^16, 17^ Available studies have demonstrated that impaired renal function, as assessed by serum levels of creatinine^10, 18–20^ or CysC^10, 18, 21–25^, predicts future CVD. However, most of these studies were conducted in non-pregnant older women, and limited data are available to address the role of these renal biomarkers measured shortly after delivery on severe maternal CVD morbidities later in life, individually and in combination with pregnancy complications.

The goal of the present study was to investigate the impacts of immediate postpartum creatinine and CysC on the mothers’ later development of CVD, using data from the Boston Birth Cohort (BBC), a long-running, predominantly urban, racially diverse, and low-income birth cohort. We hypothesized that elevated levels of these postpartum renal biomarkers, in combination with pregnancy complications, were significantly associated with an increased risk of maternal CVD risk.

## Methods

This study adheres to the American Heart Journals’ implementation of the transparency and Openness Promotion Guidelines. The data are not publicly available because the parent study is still ongoing. Requests to access the dataset may be sent to Dr Xiaobin Wang at the Johns Hopkins Bloomberg School of Public Health and will be reviewed by the investigative team and the Johns Hopkins Bloomberg School of Public Health institutional review board (IRB).

### Study Design and Population

The BBC was initiated in 1998 as a prospective birth cohort, with continued follow up of mother-child dyads at the Boston Medical Center^26, 27^. Women admitted to the labor and delivery floor who delivered a singleton live infant without major birth defects were eligible for participation in this cohort. Research staff approached and recruited mothers 24 to 72 hours after delivery. Mothers were enrolled beginning in 1998 and were followed until 2019. We obtained written informed consent from each mother in the parent study. The Boston Medical Center and the Johns Hopkins Bloomberg School of Public Health IRBs approved this study. The study is also registered on ClinicalTrials.gov (NCT03228875).

In the BBC, 2,874 independent mother-infant pairs were followed until 2016, with a follow-up rate of ∼85%. For this study, 583 mothers were randomly selected for measurement of renal markers, and these samples were enriched for mothers with preeclampsia (with a ratio of ∼1:3 for those with and without preeclampsia). Seven mothers with chronic kidney disease (CKD) before delivery, as documented in maternal electronic medical records, were excluded from analyses. **Table S1** presents population characteristics for the 576 selected mothers compared to mothers not selected in this study, indicating that these two groups of mothers were comparable on population characteristics including maternal marital status, parity, perceived stress during pregnancy, and smoking during pregnancy, and on incident rate of CVD.

### Measurement of immediate postpartum creatinine, CysC and eGFR calculation

Maternal blood samples were collected 24 to 72 hours after delivery. EDTA plasma was separated from red blood cells by centrifugation, and all the samples were stored at −80℃. Plasma was aliquoted, randomly distributed into each plate, and sent to the Clinical Chemistry Research Laboratory, University of Maryland, Baltimore, for biomarker measurement in Oct 2021. Creatinine was measured using the ECREA Flex reagent cartridge on the Dimension Vista System (Siemens Healthcare Diagnostics, Malvern, PA), and cystatin C (CysC) was measured using the CYSC method on the Dimension Vista System. The laboratory ran quality control (QC) daily and assured that the QC results were within acceptable ranges. Besides, we measured seven duplicate samples on different plates. The mean inter-assay coefficient of variation for these duplicates was 5.9% for creatinine and 6.6% for CysC. Calculations of eGFR were made using the most recent creatinine- and CysC-based equation incorporating age and sex parameters.^28^

### Diagnosis of preeclampsia and preterm delivery

Data on physician-diagnosed preeclampsia and eclampsia were extracted from maternal electronic medical records (EMR), as described^29^. For women with preexisting chronic hypertension, we required evidence of worsening hypertension (defined as systolic blood pressure ≥160 mmHg and/or diastolic blood pressure of ≥110 mmHg). The syndrome of hemolysis, elevated liver enzymes, and low platelets developing during pregnancy (HELLP) was considered present when a physician made this diagnosis based on chart review. We combined cases of eclampsia and HELLP with preeclampsia because there were too few affected women to allow for separate groups. All relevant medical records of preeclampsia cases were reviewed to ensure that they met the definition of preeclampsia according to the recent American College of Obstetricians and Gynecologists criteria^30^.

Gestational age was assessed by early prenatal ultrasound (<20 weeks) or if early prenatal ultrasound was not available, was based on the first day of the last menstrual period as recorded in the maternal EMR.^26^ Preterm delivery was defined as gestational age <37 weeks. For the degree of preterm delivery, we defined early preterm delivery as gestational age <32 weeks, and late preterm delivery as gestational age within 32-36.9 weeks.

### Diagnosis of severe maternal CVD

Maternal diagnosis of severe CVD (referred as “CVD”) was based on ICD-9-CM codes (for diagnosis before or on Oct 1, 2015) and ICD-10-CM codes (for diagnosis after Oct 1, 2015), extracted from maternal annual EMR (until the end of 2021), as summarized in **Table S2**. The following incident phenotypic outcomes were included: 1) atrial fibrillation (AF), 2) heart failure (HF), 3) ischemic heart disease (IHD), 4) peripheral vascular disease (PVD), and 5) stroke. Women with at least one occurrence of any of these diseases (**Table S2**) were coded as having CVD.

### Covariates

A standard questionnaire interview was administered 24 to 72 hours after delivery. This collected data on maternal epidemiological factors, including self-reported race/ethnicity (non-Hispanic Black; non-Hispanic White; Hispanic; and others), birthplace (foreign-born vs US-born), education level, parity, tobacco smoking, drug use during pregnancy (including cannabis, cocaine, crack cocaine, amphetamines, heroin, methadone, barbiturates, etc.), self-reported stress during pregnancy, and alcohol consumption during pregnancy. Maternal pre-pregnancy body mass index (BMI), calculated as self-reported weight (kg) divided by height squared (m^2^), was categorized into four groups: normal weight (<25.0 kg/m^2^), overweight (25-29.9 kg/m^2^), obese (≥30 kg/m^2^), and unknown. Clinical complications before and during pregnancy, including pregestational chronic hypertension, pregestational or gestational diabetes, delivery type, and fetal sex were extracted from the EMR.

### Statistical analyses

The distribution of plasma creatinine levels (mg/dL) and CysC (mg/L) was plotted for all enrolled women, both of which were approximate to a normal distribution, but with a few extreme values or outliers at the right end. To minimize the potential impact of outliers, we first defined outliers as values more extreme than the mean plus or minus the standard deviation (SD) multiplied by 3. We identified 3 women with creatinine > mean + 3SD and 5 women with CysC > mean + 3SD. These outliers were then recoded as mean+3SD for subsequent analyses. Locally weighted scatterplot smoothing (LOESS) plots were generated to explore relationships of plasma creatinine and CysC levels with gestational age at delivery in mothers with preeclampsia and those without preeclampsia, separately. Linear regression models were applied to examine associations of a series of maternal personal and social characteristics with creatinine, CysC and eGFR, respectively.

To analyze the relationships of creatinine, CysC and eGFR with risk of CVD later in life, Cox proportional hazards models were applied to generate hazard ratios (HR) with time-to-CVD as the outcome, with years of follow-up since delivery as the time scale. We followed women from about two months after delivery until date of CVD diagnosis or until date of last visit, whichever came first. Creatinine and CysC were analyzed as both continuous and categorical exposures (quartiles). Based on the results using quartiles, we defined normal (or Q1-Q3) vs elevated (or Q4) level of creatinine and CysC as ≤ 75^th^ vs >75^th^ percentiles. eGFR was classified as both a continuous, categorical (< 60, 60-89 and ≥90 ml/min/1.73m^2^, reflecting kidney damage, mildly reduced kidney function, and normal kidney function, respectively) and binary exposure (< 60 vs ≥60 ml/min/1.73m^2^). For each Cox proportional hazards model, the proportional hazards (PH) assumption was assessed based on the scaled Schoenfeld residual, using R package cox.zph, indicating that the assumption was not violated in this study.

The following maternal variables were adjusted in the model based on *a priori* information from the literature: maternal age at enrollment, self-reported race/ethnicity, educational level, maternal smoking during pregnancy, maternal pre-pregnancy BMI, pregestational and gestational diabetes, pregestational chronic hypertension, preeclampsia and degree of preterm delivery. The combined effects of each renal biomarker and pregnancy complication (preeclampsia and/or preterm delivery) on time-to-CVD were analyzed using proportional hazards models, and their interactive effects were analyzed by adding their product term into the Cox proportional hazards models. We considered a two-sided *P*<0.05 as statistical significance cutoff. All analyses were performed in R-studio (Version 1.2).

## Results

### Descriptive Results

Of the 576 mothers enrolled at delivery in this study, mean age at enrollment was 29.9 (SD: 6.6) years; 364 (63.2%) were non-Hispanic Black; 322 (55.9%) were overweight or obese (OWO) before pregnancy; 90 (15.6%) had chronic hypertension; 149 (25.6%) had preeclampsia; and 177 (30.7%) had preterm delivery (**Table 1**).

**Table 1.**
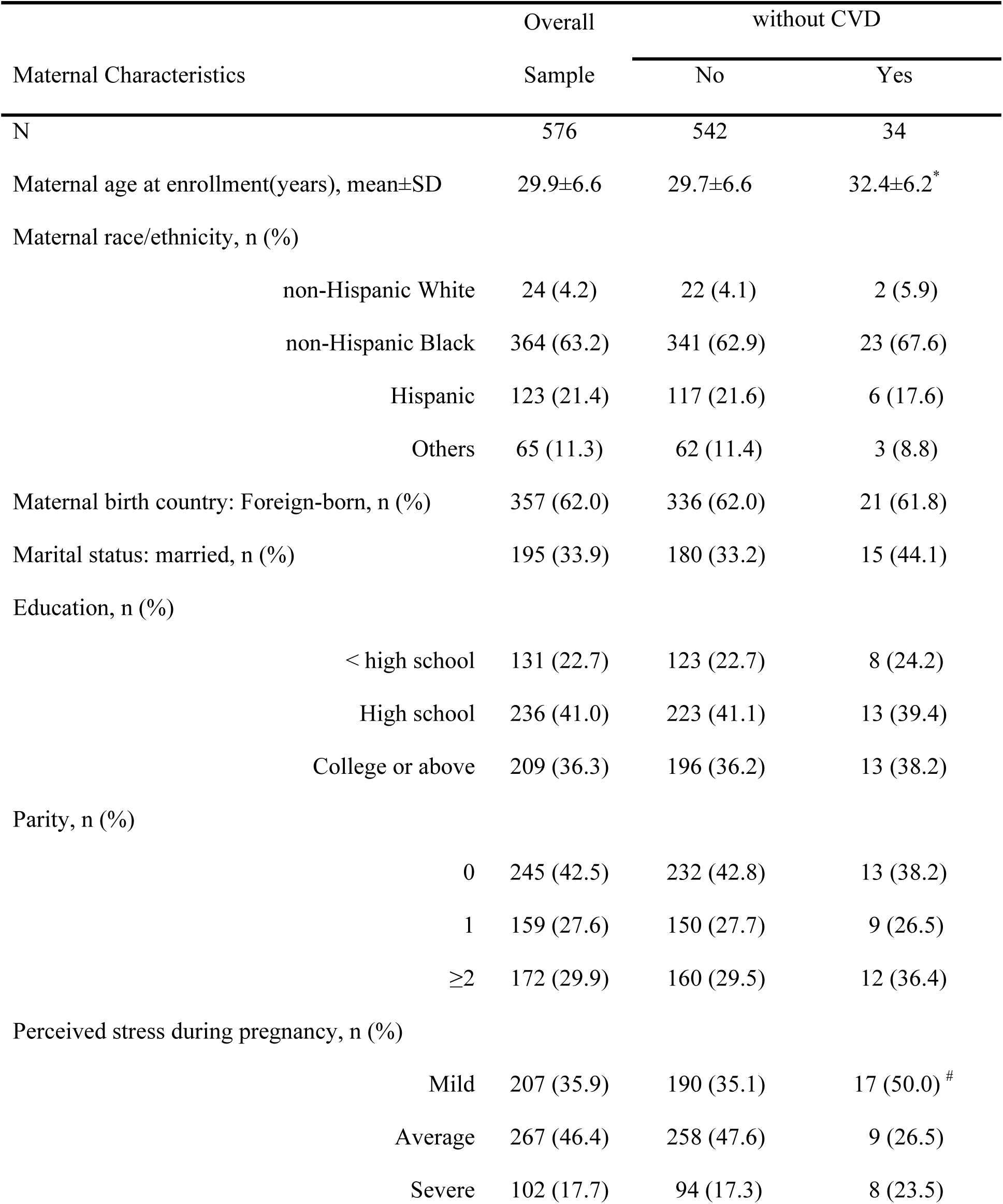
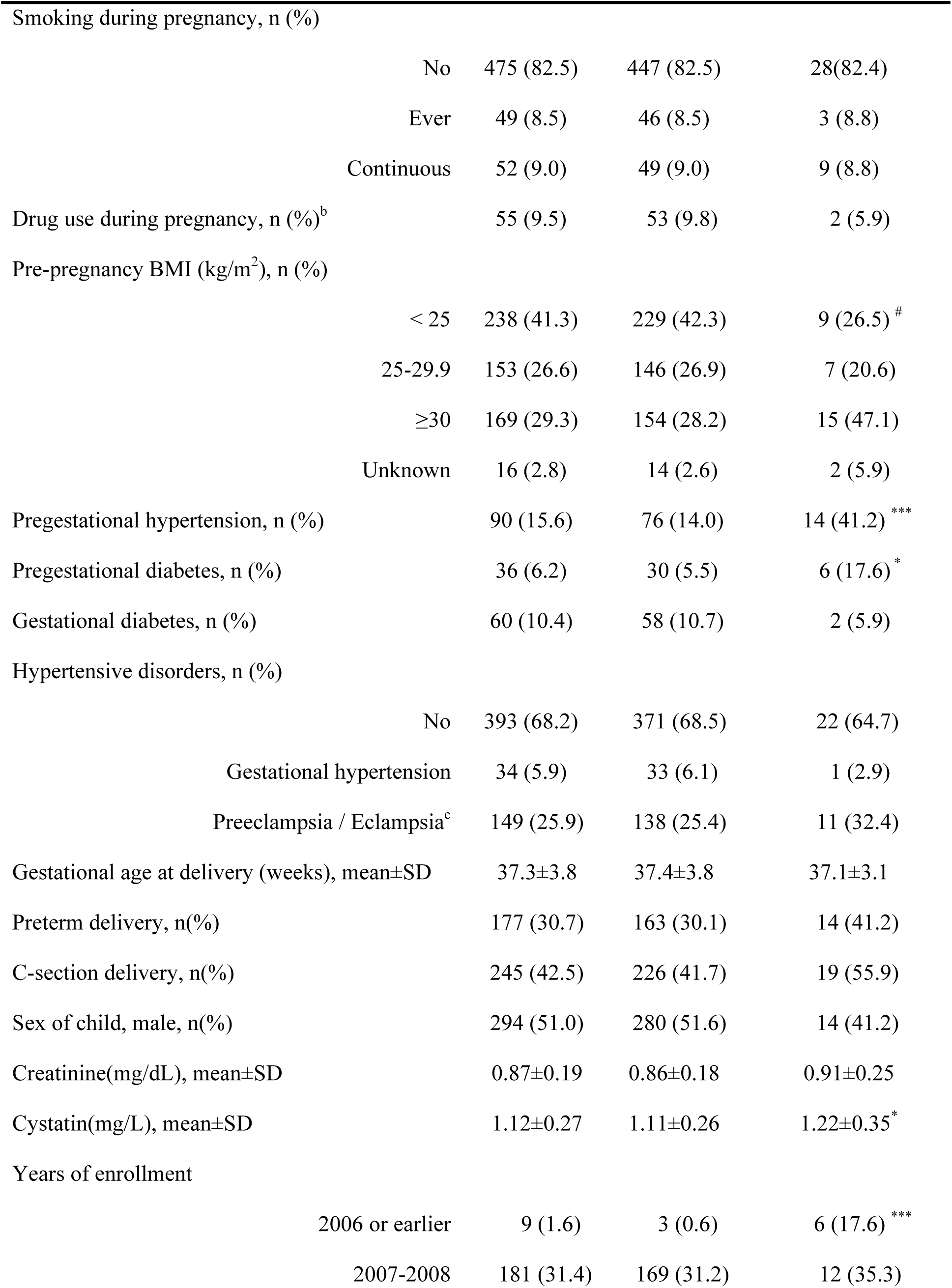
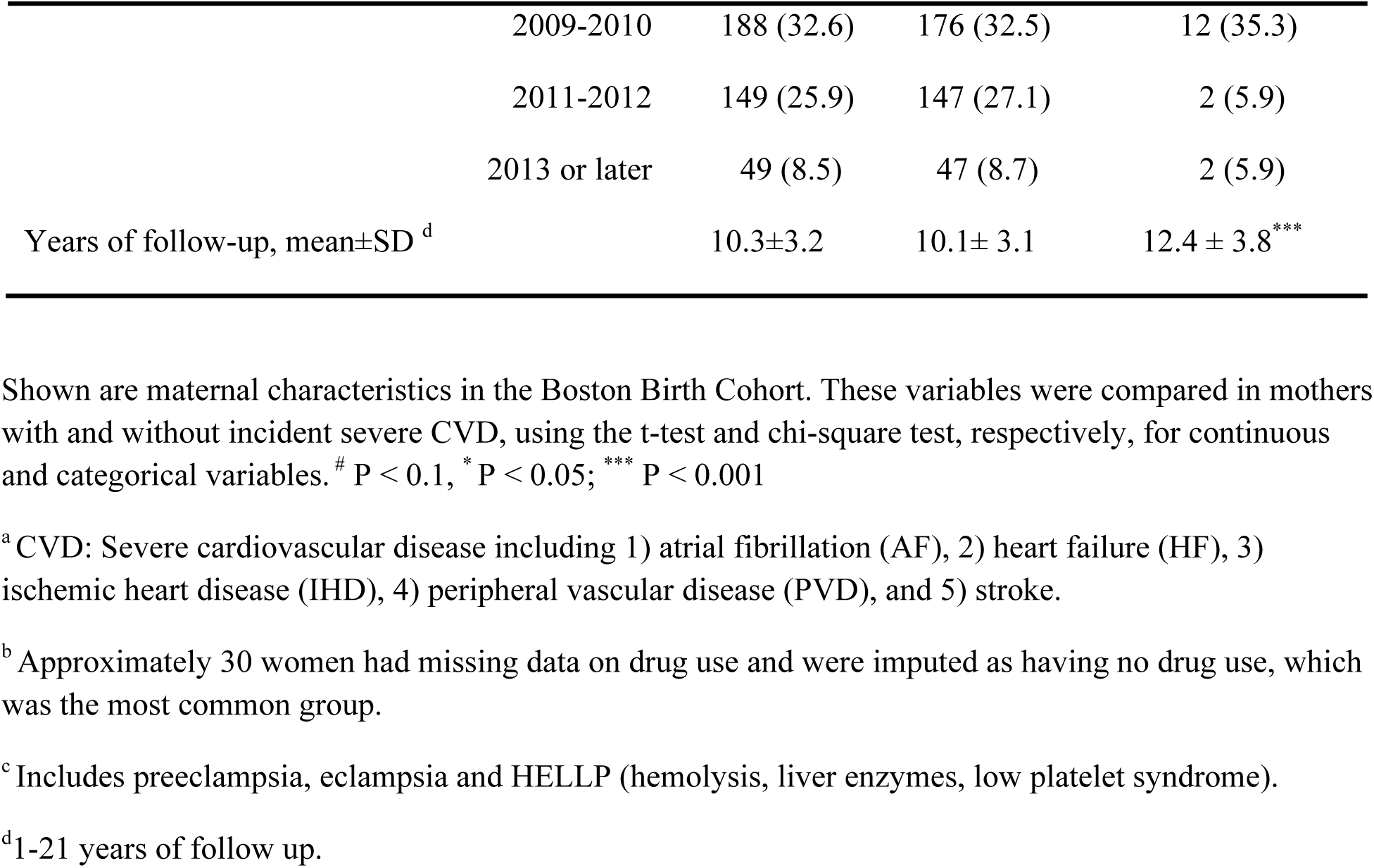
Baseline characteristics of the 576 women from the Boston Birth Cohort, stratified by incident severe cardiovascular disease (CVD) status during the postnatal follow-up.

| Maternal Characteristics | Overall | without CVD |  |
| --- | --- | --- | --- |
|  | Sample | No | Yes |
| N | 576 | 542 | 34 |
| Maternal age at enrollment(years), mean±SD | 29.9±6.6 | 29.7±6.6 | 32.4±6.2* |
| Maternal race/ethnicity, n (%) |  |  |  |
| non-Hispanic White | 24 (4.2) | 22 (4.1) | 2 (5.9) |
| non-Hispanic Black | 364 (63.2) | 341 (62.9) | 23 (67.6) |
| Hispanic | 123 (21.4) | 117 (21.6) | 6 (17.6) |
| Others | 65 (11.3) | 62 (11.4) | 3 (8.8) |
| Maternal birth country: Foreign-born, n (%) | 357 (62.0) | 336 (62.0) | 21 (61.8) |
| Marital status: married, n (%) | 195 (33.9) | 180 (33.2) | 15 (44.1) |
| Education, n (%) |  |  |  |
| < high school | 131 (22.7) | 123 (22.7) | 8 (24.2) |
| High school | 236 (41.0) | 223 (41.1) | 13 (39.4) |
| College or above | 209 (36.3) | 196 (36.2) | 13 (38.2) |
| Parity, n (%) |  |  |  |
| 0 | 245 (42.5) | 232 (42.8) | 13 (38.2) |
| 1 | 159 (27.6) | 150 (27.7) | 9 (26.5) |
| ≥2 | 172 (29.9) | 160 (29.5) | 12 (36.4) |
| Perceived stress during pregnancy, n (%) |  |  |  |
| Mild | 207 (35.9) | 190 (35.1) | 17 (50.0) # |
| Average | 267 (46.4) | 258 (47.6) | 9 (26.5) |
| Severe | 102 (17.7) | 94 (17.3) | 8 (23.5) |

|  |  |  |  |  |
| --- | --- | --- | --- | --- |
| Smoking during pregnancy, n (%) |  |  |  |  |
|  | No | 475 (82.5) | 447 (82.5) | 28(82.4) |
|  | Ever | 49 (8.5) | 46 (8.5) | 3 (8.8) |
|  | Continuous | 52 (9.0) | 49 (9.0) | 9 (8.8) |
| Drug use during pregnancy, n (%)<sup>b</sup> |  | 55 (9.5) | 53 (9.8) | 2 (5.9) |
| Pre-pregnancy BMI (kg/m<sup>2</sup>), n (%) |  |  |  |  |
|  | < 25 | 238 (41.3) | 229 (42.3) | 9 (26.5) <sup>#</sup> |
|  | 25-29.9 | 153 (26.6) | 146 (26.9) | 7 (20.6) |
|  | ≥30 | 169 (29.3) | 154 (28.2) | 15 (47.1) |
|  | Unknown | 16 (2.8) | 14 (2.6) | 2 (5.9) |
| Pregestational hypertension, n (%) |  | 90 (15.6) | 76 (14.0) | 14 (41.2) <sup>***</sup> |
| Pregestational diabetes, n (%) |  | 36 (6.2) | 30 (5.5) | 6 (17.6) <sup>*</sup> |
| Gestational diabetes, n (%) |  | 60 (10.4) | 58 (10.7) | 2 (5.9) |
| Hypertensive disorders, n (%) |  |  |  |  |
|  | No | 393 (68.2) | 371 (68.5) | 22 (64.7) |
|  | Gestational hypertension | 34 (5.9) | 33 (6.1) | 1 (2.9) |
|  | Preeclampsia / Eclampsia<sup>c</sup> | 149 (25.9) | 138 (25.4) | 11 (32.4) |
| Gestational age at delivery (weeks), mean±SD |  | 37.3±3.8 | 37.4±3.8 | 37.1±3.1 |
| Preterm delivery, n(%) |  | 177 (30.7) | 163 (30.1) | 14 (41.2) |
| C-section delivery, n(%) |  | 245 (42.5) | 226 (41.7) | 19 (55.9) |
| Sex of child, male, n(%) |  | 294 (51.0) | 280 (51.6) | 14 (41.2) |
| Creatinine(mg/dL), mean±SD |  | 0.87±0.19 | 0.86±0.18 | 0.91±0.25 |
| Cystatin(mg/L), mean±SD |  | 1.12±0.27 | 1.11±0.26 | 1.22±0.35<sup>*</sup> |
| Years of enrollment |  |  |  |  |
|  | 2006 or earlier | 9 (1.6) | 3 (0.6) | 6 (17.6) <sup>***</sup> |
|  | 2007-2008 | 181 (31.4) | 169 (31.2) | 12 (35.3) |
|  | 2009-2010 | 188 (32.6) | 176 (32.5) | 12 (35.3) |
|  | 2011-2012 | 149 (25.9) | 147 (27.1) | 2 (5.9) |
|  | 2013 or later | 49 (8.5) | 47 (8.7) | 2 (5.9) |
| Years of follow-up, mean±SD <sup>d</sup> |  | 10.3±3.2 | 10.1± 3.1 | 12.4 ± 3.8<sup>***</sup> |

Shown are maternal characteristics in the Boston Birth Cohort. These variables were compared in mothers with and without incident severe CVD, using the t-test and chi-square test, respectively, for continuous and categorical variables. # P < 0.1, \* P < 0.05; \*\*\* P < 0.001
<sup>a</sup> CVD: Severe cardiovascular disease including 1) atrial fibrillation (AF), 2) heart failure (HF), 3) ischemic heart disease (IHD), 4) peripheral vascular disease (PVD), and 5) stroke.
<sup>b</sup> Approximately 30 women had missing data on drug use and were imputed as having no drug use, which was the most common group.
<sup>c</sup> Includes preeclampsia, eclampsia and HELLP (hemolysis, liver enzymes, low platelet syndrome).
<sup>d</sup> 1-21 years of follow up.

During an average of 10.3 years of follow-up, severe CVD was newly diagnosed in 34 mothers (5.9%), including AF in 4 mothers, IHD in 18 mothers, HF in 7 mothers, PVD in 5 mothers and stroke in 9 mothers (**Table S2**). Compared to mothers without CVD, those who developed CVD were older at enrollment (*P* =0.02), were more likely to have pregestational hypertension (*P*< 0.001) and pregestational diabetes (*P*=0.015); They also had higher postpartum CysC levels (*P*=0.015) and longer follow-up (*P* < 0.001, **Table 1**).

### Maternal factors associated with immediate postpartum creatinine and CysC levels

**Figure S1** presents the distribution of maternal plasma creatinine and CysC values obtained 24-72 hours postpartum, with a median (inter-quartile range) of 0.83 (0.24) mg/dL and 1.09 (0.33) mg/L, respectively. **Fig 1** presents the relationships of both biomarkers with gestational age at delivery, while simultaneously considering the status of preeclampsia. Plasma creatinine levels tended to be higher with greater gestational age at delivery in non-preeclamptic mothers but not in preeclamptic mothers. In comparison, postpartum plasma CysC levels increased with greater gestational age at delivery in mothers with and without preeclampsia. Conditioned on gestational age, levels of both creatinine and CysC were higher in preeclamptic women than in non-preeclamptic women. With multivariable-adjusted linear regression models to analyze all potential maternal factors, we found that preeclampsia was significantly associated with higher plasma levels of both creatinine and CysC; on the other hand, preterm delivery (<32 weeks and 32-36.9 weeks compared to ≥37 weeks) was significantly associated with lower plasma levels of both biomarkers, with a dose-response relationship between degree of preterm delivery and CysC levels (**Table 2**). Mothers with pregestational chronic hypertension had lower CysC levels while mothers with drug use during pregnancy had higher CysC levels than those without these conditions. When eGFR was analyzed as the outcome, maternal age at enrollment, drug use during pregnancy and the existence of preeclampsia were inversely associated with eGFR, while pregestational chronic hypertension and preterm delivery were positively associated with eGFR (**Table 2**).

**Figure 1.**
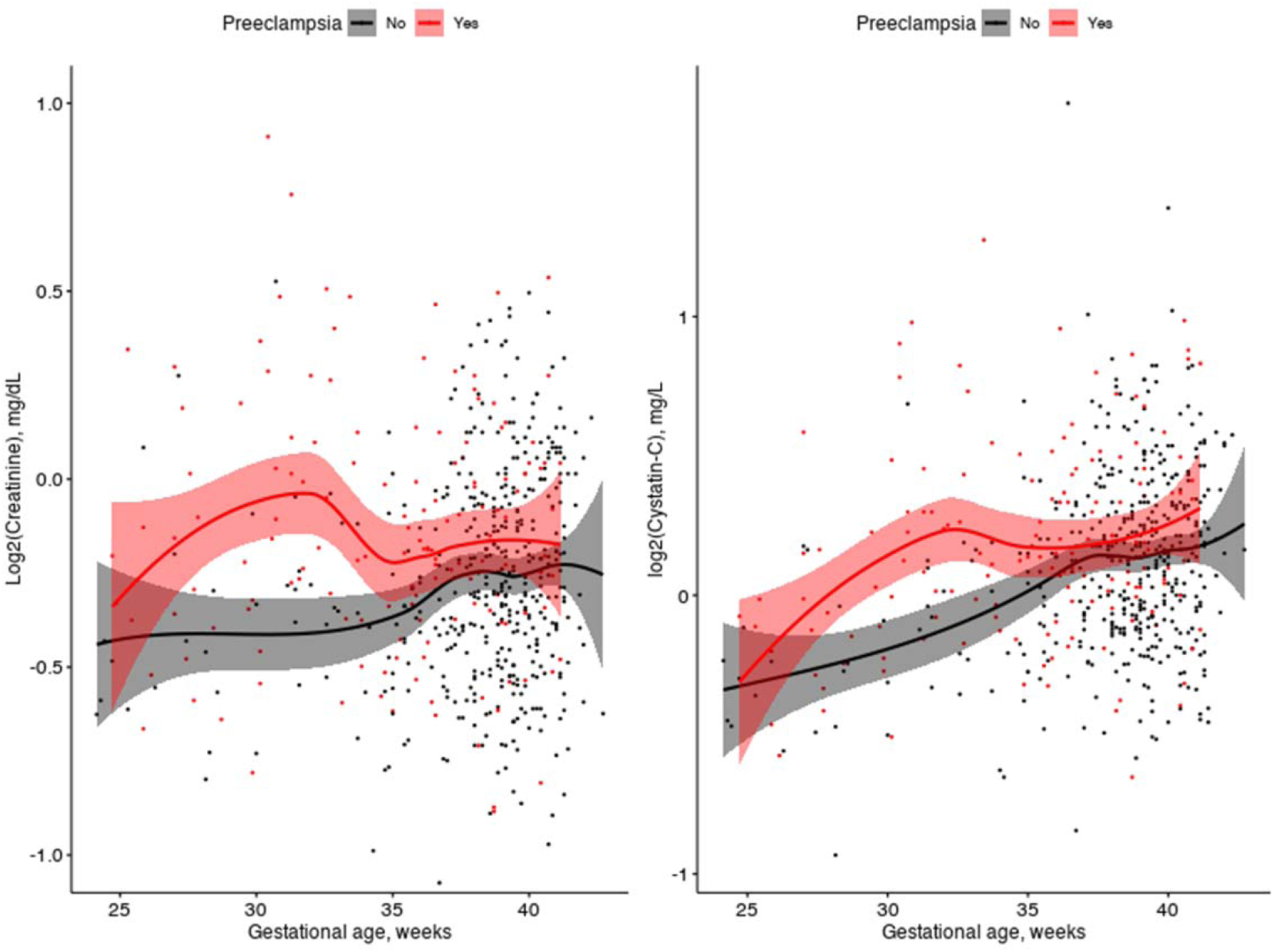
Relationships of maternal postpartum plasma creatinine and cystatin C levels with gestational age at delivery, stratified by preeclampsia status. In each graph, the y-axis is creatinine and cystatin C, respectively, the x-axis is gestational age at delivery, the line is fit with LOESS regression, and the shaded area indicates the 95% confidence interval, stratified by preeclampsia.

**Table 2.** Prenatal and perinatal determinants of maternal immediate postpartum creatinine, cystatin C and eGFR levels among 576 women.

| Maternal Variables |  | Creatinine<sup>a</sup> | Cystatin C<sup>a</sup> | eGFR<sup>a,b</sup> |
| --- | --- | --- | --- | --- |
|  |  | Beta ± SE | Beta ± SE | Beta ± SE |
| Maternal age at enrollment (years) |  | 0.00±0.00 | 0.00±0.00 | -0.49±0.16** |
| Race/ethnicity | non-Hispanic White | Ref | Ref | Ref |
|  | non-Hispanic Black | 0.05±0.04 | -0.01±0.06 | -1.71±4.37 |
|  | Hispanic | -0.02±0.04 | -0.09±0.07 | 5.89±4.84 |
|  | Other | -0.05±0.05 | -0.11±0.07 | 7.73±4.94 |
| Marital status: | Married vs unmarried | 0.00±0.02 | -0.05±0.03* | 2.76±1.96 |
| Maternal birth country: Foreign-born vs U.S.-born |  | 0.01±0.02 | 0.05±0.03 | -3.43±2.29 |
| Education level. | < High school | Ref | Ref | Ref |
|  | High school | -0.00±0.02 | -0.03±0.03 | 1.15±2.28 |
|  | College or above | 0.04±0.02 | 0.02±0.03 | -3.08±2.41 |
| Parity | 0 | Ref | Ref | Ref |
|  | 1 | -0.03±0.02 | -0.02±0.03 | 1.75±2.11 |
|  | ≥2 | -0.03±0.02 | -0.02±0.03 | 2.01±2.33 |
| Stress during pregnancy | Mild | Ref | Ref | Ref |
|  | Average | -0.02±0.02 | -0.03±0.03 | 2.08±1.91 |
|  | Severe | -0.01±0.02 | -0.03±0.03 | 1.75±2.49 |
| Smoking during pregnancy | Never | Ref | Ref | Ref |
|  | Quitter | -0.01±0.03 | 0.01±0.04 | -0.01±3.17 |
|  | Continuous | 0.03±0.03 | 0.05±0.04 | -3.97±3.28 |
| Drug use during pregnancy |  | 0.05±0.03 | 0.08±0.04* | -6.45±3.12* |
| Pre-pregnancy BMI, kg/m<sup>2</sup> | <25 | Ref | Ref | Ref |
|  | 25-29.9 | 0.02±0.02 | -0.02±0.03 | 0.45±2.10 |
|  | ≥30 | 0.01±0.02 | -0.03±0.03 | 0.36±2.12 |
| Pregestational chronic hypertension |  | -0.04±0.02 | -0.08±0.03<sup>*</sup> | 5.84±2.47<sup>*</sup> |
| Gestational hypertension |  | -0.03±0.03 | -0.03±0.05 | 4.89±3.63 |
| Preeclampsia |  | 0.09±0.02<sup>***</sup> | 0.12±0.03<sup>***</sup> | -9.89±2.10<sup>***</sup> |
| Diabetes |  |  |  |  |
|  | None | Ref | Ref | Ref |
|  | Gestational | -0.01±0.03 | 0.02±0.04 | -0.39±2.76 |
|  | Pre-gestational | 0.03±0.03 | 0.05±0.05 | -3.43±3.58 |
| Gestational age at delivery | ≥ 37 weeks | Ref | Ref | Ref |
|  | 32-36.9 weeks | -0.06±0.02<sup>**</sup> | -0.09±0.03<sup>**</sup> | 7.46±2.22<sup>***</sup> |
|  | < 32 weeks | -0.04±0.03 | -0.21±0.04<sup>***</sup> | 15.40±2.84<sup>***</sup> |
| Fetal sex: male |  | -0.01±0.02 | -0.01±0.02 | 0.77±1.66 |

**Legend:** <sup>a</sup> Shown are the beta coefficients and standard errors from the linear regression models with creatinine, cystatin C, or eGFR as the outcomes. <sup>\*</sup> $P < 0.05$ ; <sup>\*\*</sup> $P < 0.01$ , <sup>\*\*\*</sup> $P < 0.001$ . All the variables in the table were analyzed in the same linear regression model to adjust for each other covariate.
<sup>b</sup> eGFR was calculated using the creatinine- and cystatin C based equation including coefficients for age and sex: $eGFR = 135 \times \min(Cr/0.7, 1)^{-0.219} \times \max(Cr/0.7, 1)^{-0.544} \times \min(CysC/0.8, 1)^{-0.323} \times \max(CysC/0.8, 1)^{-0.778} \times 0.9961^{age} \times 0.963$ . Cr: Creatinine in mg/dL; CysC: cystatin C in mg/L.

### Maternal postpartum renal biomarkers and maternal risk of CVD during follow-up

There was no statistically significant association between creatinine and CVD risk (**Table 3**). By contrast, per unit increase of CysC was associated with a 5.2-fold higher risk of developing CVD (95% CI=1.5-18.2, *P*=0.010). When CysC quartile was analyzed as a categorical variable and with women in the lowest quartile (Q1) as the reference, those in the 2^nd^ (Q2) or in the 3^rd^ (Q3) quartile shown no significant difference in CVD risk; while those in the highest CysC quartile (Q4) were at a significant higher risk of developing CVD (HR=3.5, 95%CI=1.1-11.3, *P*=0.04). We therefore defined elevated (or Q4) vs normal (or Q1-Q3) CysC with the 75^th^ percentile (1.26 mg/L) as the cutoff. Population characteristics between these two groups were shown in **Table S3**.

**Table 3.**
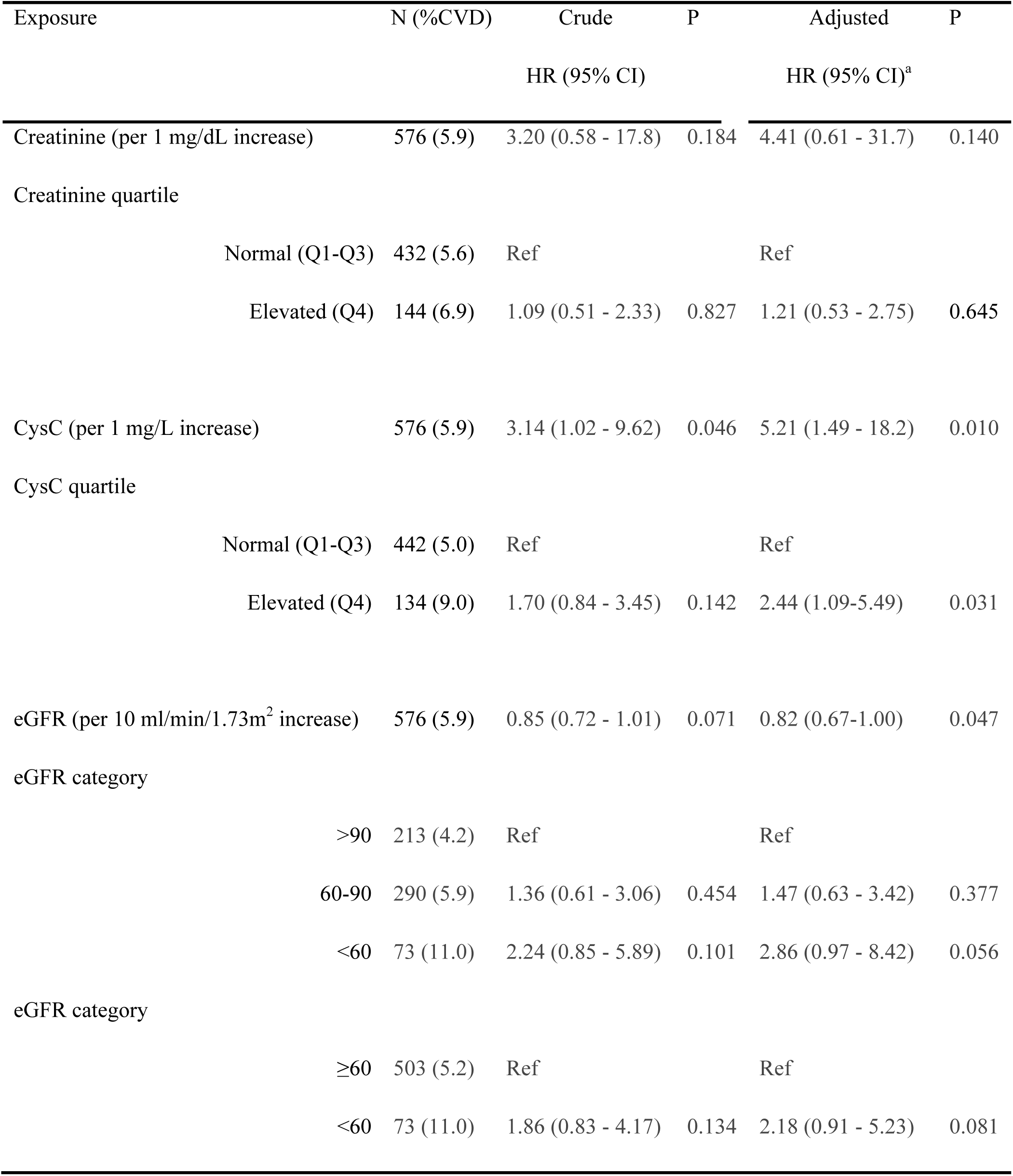
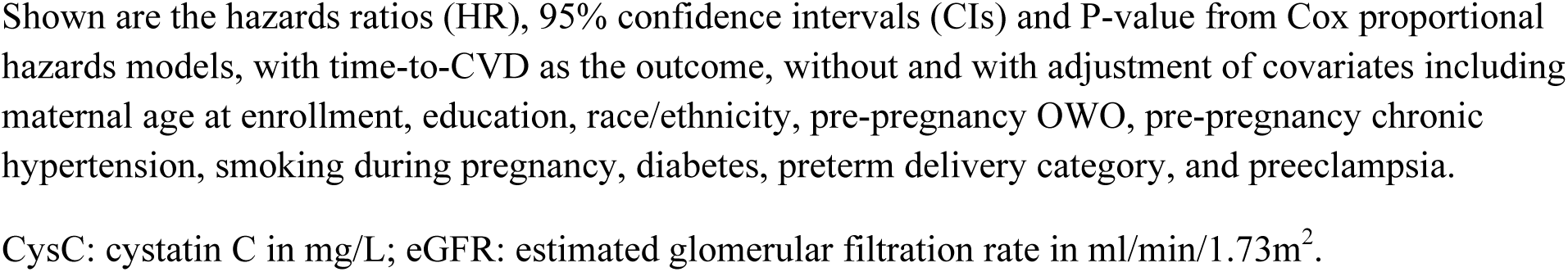
Associations (Hazard ratio and 95% CI) of maternal cystatin C, creatinine and estimated glomerular filtration rate with incident cardiovascular diseases (CVD) among 576 women.

**Figure S2** presents the incidence rate of overall CVD and each CVD event in mothers with elevated vs normal CysC. With adjustment of covariates, we found that elevated CysC was associated with an HR of 2.4 (95%CI=1.1-5.5, P=0.031) in predicting CVD risk (**Table 3**), compared to those with normal CysC (≤75th percentile). The positive association remained largely unchanged after adjusting for creatinine in the model (data not shown). With eGFR analyzed as the exposure, mothers with postpartum eGFR< 60 ml/min/1.73m^2^ had an HR of 2.9 (95% CI=1.0-8.4, p=0.056) in association with CVD risk later in life, compared to those with eGFR ≥ 90 ml/min/1.73m^2^. Similar findings were observed when mothers with eGFR between 60 and 90 ml/min/1.73m^2^ and mothers with eGFR > 90 ml/min/1.73m^2^ were combined into a single group, serving as the reference (**Table 3**). These results remained comparable with the adjustment of gestational age at delivery as a continuous variable instead of degree of preterm delivery (data not shown), or after removing 10 mothers who likely have had a CVD event before pregnancy (data not shown), or after removing 90 mothers with pregestational chronic hypertension (**Table S4**).

### The combined effects of maternal postpartum complications and pregnancy complication

As presented in **Table 4**, with non-preeclamptic women having normal CysC values (Q1-Q3) as the reference group, mothers with preeclampsia only or mothers with elevated CysC (Q4) only showed no significant difference in CVD risk. On the other hand, mothers with both preeclampsia and elevated CysC were at the highest risk of developing CVD (HR=3.8, 95%CI=1.4 - 10.2, *P*=0.008). There was a borderline significant interaction between CysC and preeclampsia in association with CVD risk (P for interaction =0.064). Similar combined effects were observed between CysC and preterm delivery, with those women having preterm delivery and elevated CysC at the highest risk of developing CVD (HR=5.9, 95%CI=1.8-19.8, P=0.004, **Table 4**).

**Table 4.** Combined associations (hazard ratio and 95% CI) of maternal cystatin C and pregnancy complications with incident cardiovascular diseases (CVD) among 576 women.

| Pregnancy complication | CysC<sup>a</sup> | n | %CVD | crude HR | P | Adjusted HR<sup>b</sup> | P |
| --- | --- | --- | --- | --- | --- | --- | --- |
| **Preeclampsia as pregnancy complication** |  |  |  |  |  |  |  |
| No | Normal | 336 | 5.1 | Ref |  | Ref |  |
| No | Elevated | 91 | 6.6 | 1.15 (0.45 - 2.95) | 0.771 | 1.22 (0.44 - 3.34) | 0.704 |
| Yes | Normal | 106 | 4.7 | 0.98 (0.26 - 2.07) | 0.963 | 0.71 (0.25 - 1.99) | 0.512 |
| Yes | Elevated | 43 | 14.0 | 3.01 (1.18 - 7.68) | 0.020 | 3.81 (1.43 - 10.2) | 0.008 |
| Interaction |  |  |  |  | 0.196 |  | 0.064 |
| **Preterm delivery as pregnancy complication** |  |  |  |  |  |  |  |
| No | Normal | 291 | 4.1 | Ref |  | Ref |  |
| No | Elevated | 108 | 7.4 | 1.61 (0.65 - 3.96) | 0.297 | 1.84 (0.70 - 4.82) | 0.214 |
| Yes | Normal | 151 | 6.6 | 1.55 (0.67 - 3.59) | 0.306 | 1.44 (0.60 - 3.42) | 0.413 |
| Yes | Elevated | 26 | 15.4 | 4.12 (1.33 - 12.8) | 0.014 | 5.89 (1.75 - 19.8) | 0.004 |
| Interaction |  |  |  |  | 0.505 |  | 0.308 |
| **Preeclampsia and preterm delivery as pregnancy complications** |  |  |  |  |  |  |  |
| Neither | Normal | 254 | 3.5 | — |  | — |  |
| Neither | Elevated | 86 | 5.8 | 1.26 (0.42 - 3.73) | 0.679 | 1.32 (0.42 - 4.16) | 0.636 |
| Either | Normal | 119 | 7.6 | 1.86 (0.74 - 4.59) | 0.178 | 1.82 (0.72 - 4.58) | 0.204 |
| Either | Elevated | 27 | 14.8 | 4.22 (1.32 - 13.5) | 0.015 | 5.82 (1.70 - 19.9) | 0.005 |
| Both | Normal | 69 | 4.3 | 1.12 (0.31 - 4.08) | 0.862 | 0.81 (0.21 - 3.08) | 0.754 |
| Both | Elevated | 21 | 14.3 | 3.93 (1.08 - 14.3) | 0.038 | 5.21 (1.34- 20.2) | 0.017 |
| Interaction |  |  |  |  | 0.311 |  | 0.125 |

**Table legend:** Shown are the hazard ratios (HR), 95% confidence intervals (CIs) and P-value from Cox proportional hazard models, with time-to-CVD as the outcome, without and with adjustment of covariates including maternal age at enrollment, education, race/ethnicity, pre-pregnancy OWO, pre-pregnancy chronic hypertension, smoking during pregnancy and diabetes.
<sup>a</sup> Elevated (Q4) vs normal (Q1-Q3) cystatin C (CysC) was defined as CysC > 75<sup>th</sup> percentile (or > 1.26mg/L) vs CysC ≤ 75<sup>th</sup> percentile.

We further classified all women into six groups based on two parameters: CysC (elevated vs normal) and number of pregnancy complication (neither preeclampsia nor preterm delivery, either complication, or both complications). With women having no pregnancy complication and having normal CysC as the reference group, we found that women who experienced one or both pregnancy complication only or women with elevated CysC only showed insignificant differences in CVD risk. Women with one pregnancy complication and with elevated CysC were at a HR of 5.8 (95%CI=1.7-19.9, p=0.005) of developing CVD, and women with both pregnancy complications and with elevated CysC were associated with CVD risk, with an HR of 5.2 (95%CI=1.3-20.2, P=0.017). Similar combined effects were observed between eGFR and pregnancy complication (**Table S6**), but not between elevated creatinine and pregnancy complication (**Table S5**).

## Discussion

In this prospective U.S urban, low-income, multi-ethnic cohort study, the major findings were as follows: 1) pregnancy complications, including preeclampsia and preterm delivery, were significantly associated with altered levels of creatinine and CysC measured within 1-3 days after delivery; 2) women with higher levels of postpartum CysC were at higher risk of developing CVD later in life, and this was independent of pregnancy complications and other maternal characteristics; 3) there were synergic effects between maternal postpartum CysC and pregnancy complications in predicting CVD, suggesting that both elevated CysC during pregnancy and pregnancy complications might be used clinically as risk factors to predict CVD development later in life.

Although the determinants of plasma creatinine and CysC levels have been well-studied, and include race/ethnicity^31–33^ and metabolic disorders^34–36^, limited studies have been focused on maternal factors during pregnancy, especially in minority populations. In pregnant women, plasma CysC was higher in pregnant women with preeclampsia than those without^37, 38^. Women with a history of preeclampsia also tended to have a higher level of postpartum CysC.^39^ Consistently, the present study of multi-ethnic women, found that mothers with preeclampsia had significantly higher levels of postpartum creatinine or CysC than mothers without preeclampsia, conditioned on gestational age. No such association was observed for gestational hypertension. A recent study also indicates that CysC level is significantly associated with preeclampsia but not with gestational hypertension.^39^ Collectively, these add further evidence to suggest CysC as a potential biomarker for preeclampsia (but not for gestational hypertension), likely due to associated renal dysfunction.

Gestational age is another important factor that may be associated with levels of renal biomarkers. A previous study reported that creatinine increased steadily after 32 weeks’ gestation till a few weeks postapartum.^9^ In a total of 175 women, Schmitz et al measured renal markers at each trimester and at 4-13 weeks postpartum. They reported that both creatinine and CysC levels were highly dynamic and increased during pregnancy.^40^ In the present study, we found that gestational age at delivery was positively associated with postpartum creatinine only in women without preeclampsia (but not in women with preeclampsia); and greater gestational age was significantly and positively associated with postpartum CysC levels in both women with and without preeclampsia, although the mechanisms are largely unknown.

Previous population-based studies have shown that elevated plasma CysC predicts CVD events and mortality, which were mainly conducted among cohorts of older individuals^21, 23–25^. The current study expands these findings to a younger population (aged 29.9 ± 6.6 years). In this multi-ethnic prospective cohort, we found that postpartum plasma CysC was significantly associated with an increased risk of CVD later in life. Some studies further demonstrated that the impact of CysC on CVD outcomes may be at least partly independent of renal function^25, 41, 42^. The present study support this hypothesis, as the associations between plasma CysC and CVD risk were independent of creatinine, which is consistent with the findings reported by Koenig *et al.*^21^ We further observed that CysC appears to have a stronger association with CVD risk than eGFR which is estimated based on both CysC and creatinine. It remains unclear whether there is a causal relationship between CysC and CVD risk. Alternatively, there may be shared genetic or environmental determinants (such as inflammation) underlying both CysC levels and CVD that may drive the associations between CysC and CVD as observed in this study, which await further exploration.

A novel finding of the present study is that postpartum CysC and pregnancy complications had borderline interactive (with preeclampsia) or synergistic effects (with preterm delivery) in predicting CVD risk. CysC serves as combined marker for CKD and for systemic inflammation.^43^ Thus, CysC has added value, beyond pregnancy complications, in predicting CVD development. Since the combination of pregnancy complications and elevated postpartum CysC is associated with higher CVD risk later in life, compared to women with one condition alone, it is possible that there is complex interplay between these two conditions involved in the development of CVD. It is also likely that plasma CysC, which was higher in women with preeclampsia, may partly mediate the adverse impact of preeclampsia on CVD risk, although this hypothesis needs further investigation. Our findings further raise the possibility that appropriate screening and management of CysC levels postpartum could mitigate the adverse effects of pregnancy complication on CVD risk. This hypothesis, if it can be validated in other large cohorts, may facilitate shifts in focus from treatment to prevention of CVD in this high-risk population.

The present study has several strengths. It applied a prospective birth cohort study design to avoid reverse causality underlying the CysC - CVD association. The study sample included most Black (63%) and Hispanic (21%) populations, which historically bear a disproportionate burden of CVD but have been underrepresented in cardiovascular research^44, 45^. Several limitations in the present study should be acknowledged. First, the sample size was relatively small and there was possible inflated type 1 error due to multiple testing. Further, we enriched for women with preeclampsia during data acquisition to enhance statistical power. Thus, findings from this study await validation in a larger population and caution is needed when generalizing our findings to other populations with lower prevalence of preeclampsia. Second, data on maternal creatinine and CysC levels throughout pregnancy were not available in this study, and thus temporal relationships of these biomarkers with preeclampsia and preterm delivery remain unclear. Third, the findings on the relationship between CysC and CVD risk may be confounded by other unadjusted factors, including anti-hypertensive treatment during pregnancy, and the presence of pre-existing renal dysfunction which cannot be captured via ICD-codes due to lack of early renal screening biomarkers before or during early pregnancy. Fourth, in contrast to other women’s health cohorts with older age women, this study consists of reproductive age women followed from postpartum forward. It would be of interest for future larger scale studies to discern the associations of CysC with future CVD events occurring shortly after delivery, during subsequent pregnancy, and during non-pregnant period, which may help gain additional insight into the pathophysiology.

**In summary,** in this prospective multi-ethnic cohort study, we demonstrated that plasma CysC, independently and in combination with pregnancy combination, were associated with future maternal risk of CVD. This suggests that pregnancy may be a ‘stress test’ in which factors such as pregnancy complication and postpartum elevated CysC levels serve as the early predictive biomarkers for lifetime CVD risk. Clearly, more research is needed to examine the mechanisms or pathways linking CysC levels with CVD risk in women experiencing pregnancy complications and to further investigate whether longitudinal trajectory of CysC during follow-up may more effectively predict CVD risk. These efforts may offer opportunities to improve postpartum care to prevent or mitigate maternal long-term cardiometabolic morbidities among high-risk women, especially those with pregnancy complications.

## Data Availability

This is an ongoing cohort study. To ensure confidentiality of participants, the research data are not publicly available. However, the relevant data reported in this study is available upon reasonable request and after IRB review and approval

## Acknowledgements

We thank all BBC participants for supporting this study. We are also grateful for the dedication and hard work of the field team at the Department of Pediatrics, Boston University Chobanian & Avedisian School of Medicine, and for the support of the obstetric nursing staff at Boston Medical Center. The authors thank Linda Rosen of the Boston University Clinical Data Warehouse for assistance in obtaining relevant clinical information; the Clinical Data Warehouse service is supported by Boston University Clinical and Translational Institute and the National Institutes of Health Clinical and Translational Science Award (grant U54-TR001012). We acknowledge support from the Intramural Research Program, NIDDK, NIH.

## Non-standard Abbreviations and Acronyms

AF: Atrial fibrillation
BBC: Boston Birth Cohort
BMI: Body mass index
CKD: Chronic kidney disease
CVD: Cardiovascular disease
CysC: Cystatin C
EMR: Electronic medical records
GFR: Glomerular filtration rate
HF: Heart failure
HR: Hazard ratios
IHD: Ischemic heart disease
IRB: Institutional review board
LOESS: Locally weighted scatterplot smoothing
PH: Proportional hazards
QC: Quality control
SD: Standard deviation

